# Myopia is not a global epidemic: - what can we learn from a longitudinal study conducted in Sweden?

**DOI:** 10.1101/2022.08.31.22279457

**Authors:** Pelsin Demir, Karthikeyan Baskaran, Pedro Lima Ramos, Thomas Naduvilath, Padmaja Sankaridurg, Antonio Filipe Macedo

**Affiliations:** Medicine and Optometry, Linnaeus University, Kalmar, Sweden; Brien Holden Vision Institute, Sydney, NSW, Australia; Department and Center of Physics – Optometry and Vision Science, University of Minho, Braga, Portugal

**Keywords:** incidence, myopic shift, myopia, environmental factors, parental myopia, refractive error

## Abstract

**Background:** The prevalence of myopia in Scandinavia seems to differ from other parts of the world and the reasons remain poorly investigated. The current study investigated the incidence of myopia, myopic shift, and associated risk factors in Swedish schoolchildren. This study also investigated the development of refractive error under the effect of COVID-19 restrictions.

**Methods:** This longitudinal study was conducted between Jan-2019 and June-2021 in which a cohort of Swedish schoolchildren aged 8-16 years were recruited. Myopia was defined as spherical equivalent refraction (SER) -0.50D. Myopic shift was defined as a minimum change in SER of -0.50D between each visit. Cumulative incidence (CIN) and incidence rate (IRA) were computed. Cox-regression and linear mixed models were used to modulate myopic shift and changes in SER.

**Results:** The study enrolled 128 participants, 86% Caucasian, 70 females, mean age 12.0 years (SD=2.4). The CIN of myopia during the two-years follow-up was 5.5%, IRA of myopia was 3.2 cases per 100 person-years. The CIN of myopic shift during the two-years was 21.0%, IRA of myopic shift was 12.4 cases per 100 person-years. Cox regression revealed that the probability of myopic shift reduced with *age* and increased with *axial length/corneal-curvature ratio*. Myopic children at the baseline and children with two myopic parents showed a significant faster-paced negative SER change over time. Changes in SER during the first year of the study were more marked than changes during the second year that coincided with the Covid pandemic.

**Conclusions:** In the current study the incidence of myopia and myopic shift was low when compared with countries in East Asia. Parental myopia remains a critical factor to consider when predicting myopia progression. Progression of myopia was unaffected by restrictions imposed during COVID-19 in Sweden. In addition to ethnicity, lifestyle and adequate educational pressure might be factors keeping prevalence of myopia under control in Scandinavia. Further studies to investigate these hypotheses are warranted.

**Key messages:** *What is already known on this topic:* The rate of myopia is increasing in many regions of the world, and the recent lockdowns caused by the COVID-19 pandemic has exacerbated this problem. The highest rates of myopia are from East Asia and are in sharp contrast to the low levels reported from Scandinavia. There is a lack of studies exploring the incidence of myopia in the Scandinavian population. We conducted this study to identify the incidence of myopia among Swedish children and to determine whether the COVID-19 pandemic had any significant effects.

*What this study adds:* This study is the first to report the incidence of myopia and the myopic shift in Scandinavia. The study results showed that myopia and myopic shift were low compared to other parts of the globe. Myopia progression was not affected by the pandemic, perhaps due to Sweden’s avoidance of severe restrictions during the outbreak. Parental myopia was the significant risk factor for the progression of myopia in this cohort of Swedish school children.

*How this study might affect research, practice, or policy:* It is clear from this study’s results that parental myopia is a risk factor for myopia and is an influential predictor to consider in clinical trials that evaluate interventions to slow down the progression of myopia. Outdoor lifestyle and less educational pressure at young ages may be contributing factors to the low prevalence and incidence of myopia in Swedish children.

**Synopsis:** Incidence of myopia was low in Swedish schoolchildren despite COVID-19 pandemic and the associated risk factor was parental myopia and younger age.

## Introduction

The rates of myopia in Scandinavia seem to differ from other parts of the world. A recent study from Denmark found that the prevalence of myopia has been stable in the past 140 years^1^. These findings are in line with other studies reporting 13.4% (CI95=8.7-18.3) of myopia prevalence in Norway^2^ and 10.0% (CI95= 4.4-14.9) in Sweden^3^. This is in sharp contrast with other regions of the globe where the prevalence of myopia amongst children is estimated to be between 50.0% and 71.0%^4-6^. It remains unclear which factors can explain these contrasting results in prevalence of myopia.

Currently, there is evidence that an association exist between parental myopia and the risk of myopia in a child^3,7-9^. Some studies have shown that the odds ratio of having myopia in childhood with two myopic parents is around 3 when compared to children with no parental myopia^10^.. In addition, myopic children with two myopic parents also exhibited higher myopia (−2.33 D) when compared with children whose parents were not myopic (−1.13 D). In our previous study we also found significantly more negative refraction among children with two myopic parents compared with children whose parents were not myopic^3^. Therefore, it seems that there is a genetic predisposition because parents with myopia tend to have children also with myopia. However, genetics may be only part of the picture and genetic predisposition might be “activated” or “accelerated” when individuals are exposed to environmental risk factors.

Environmental factors such as near work and outdoor time have been associated with the incidence of myopia^9,11,12^. For outdoor time, the most common result is that extended outdoor time may delay the onset of myopia^22^. There are currently conflicting results for near work. Some studies point that sustained near work is a risk factor for myopia development and progression while others have failed to confirm this association^9,11,12^

There is a lack of studies looking at the incidence of myopia and the progression of refractive errors in Swedish children. Likewise, there is a lack of studies investigating risk factors affecting myopia incidence and progression in Swedish children. Having this information is relevant to understand the prevalence of myopia in Sweden and in Scandinavia and to generate new ideas to tackle the high incidence of myopia in other parts of the globe. In addition, there are only a few studies that were able to follow children during the COVID-19 pandemic^13^. During this period many societies implemented lockdowns which can be considered a risk for myopia progression. The aim of the current study was to investigate the incidence of myopia, myopic shift, and associated risk factors in Swedish schoolchildren. Because it was partially conducted during the COVID-19 pandemic, this study also investigated the development of refractive error under the effect of COVID-19 restrictions.

## Methods

This was longitudinal study involving a cohort of 128 children conducted in southern Sweden from January 2019 to June 2021. Readers are referred to our previously paper on baseline characteristics and recruitment procedures^3^. The study protocol complied with the tenets of the Declaration of Helsinki, informed consent was obtained from participants and their parents. The study received ethics approval from the Regional Committee for Medical Research Ethics in Linköping (Dnr 2018/423-31).

The follow-up period was 2 years with a total of four visits: baseline, 0.5, 1 and 2 years. An identical comprehensive eye examination was repeated at every visit to determine changes in refractive error and changes in ocular parameters. Details of the methodology has been given in our previous publication^3^. In brief, each visit included measurements of: distance visual acuity, axial length and corneal curvature with noncontact optical coherence biometry (IOLMaster 500 (Carl Zeiss Meditec, AG, Jena, Germany, https://www.zeiss.com/corporate/int/home.html), hight, weight and cycloplegic refraction (NVision-K 5001autorefractor (Shin-Nippon, Rexxam, http://www.shin-nippon.jp/). To ensure paralysis of the ciliary muscle and complete dilation, accommodation and pupil response were verified with a RAF rule and a penlight respectively. Cycloplegic refraction was taken 30 minutes after installation of two drops of cyclopentolate 1% (Cyclogyl, Alcon, https://www.alcon.se/sv).

Questionnaires completed by parents were collected at baseline and follow-up assessments.^3^ The questions covered demographics, parental myopia, medical history, academic preferences of the child, education, living conditions, time spent on near work and time spent on outdoor activities after school, and reading habits. Parental myopia was confirmed by analysis of the parents prescription.

### Definitions and Statistical analysis

The definition of myopia was based on cycloplegic spherical equivalent refraction (SER) of the right eye: SER= sphere + cylinder/2, cylinder with negative sign. A participant with SER of -0.50 dioptre (D) or more negative was considered myopic. Hyperopia was defined as SER +0.75 D or more positive^3^. Myopic shift was defined as a change of -0.50 D or more in SER between visits, SER at 0.5 years was subtracted from SER at baseline, then SER at 1 year was subtracted from SER at 0.5 years, and SER at 2 years was subtracted from SER at 1 year. Cumulative incidence was computed by dividing the number of incident cases by the number at risk at baseline using the equation:

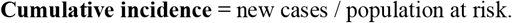

Incidence rate was computed using the equation:

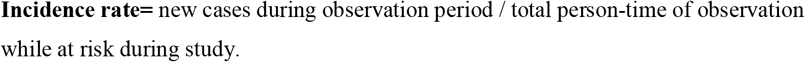

Cox proportional hazards regression model was performed to capture all possible myopic shifts (repeated events) during the 2 years of follow-up. The model included time-invariant covariates (e.g., “sex”) and time-dependent covariates (e.g., “age”) for each participant. Independent variables tested are given in results, the descriptive statistics in Table 1 and their contributions to the model in Table 2.

**Table 1.** Summary of the key variables for baseline (0), 0.5, 1, and 2 years.

|  | Baseline (n=128)<br>Mean (SD)<br>Median (IQR) | 0.5 Years (n=126)<br>Mean (SD)<br>Median (IQR) | 1 Years (n=114)<br>Mean (SD)<br>Median (IQR) | 2 Years (n=121)<br>Mean (SD)<br>Median (IQR) |
| --- | --- | --- | --- | --- |
| Age (Years) | 12.0 (SD=2.5)<br>12.0 (IQR=5.0) | 12.0 (SD=2.5)<br>12.5 (IQR=4.0) | 12.5 (SD=2.5)<br>13.0 (IQR=4.0) | 13.5 (SD=2.5)<br>14.0 (IQR=4.0) |
| SER (Diopters) | +0.65 (SD=1.2)<br>+0.70 (IQR=1.0) | +0.70 (SD=1.3)<br>+0.78 (IQR=1.1) | +0.66 (SD=1.2)<br>+0.67 (IQR=1.1S) | +0.51 (SD=1.5)<br>+0.64 (IQR=1.0) |
| AL (millimeters) | 23.0 (SD=0.86)<br>23.0 (IQR=1.1) | 23.0 (SD=0.87)<br>23.0 (IQR=1.1) | 23.0 (SD=0.82)<br>23.0 (IQR=1.1) | 23.0 (SD=0.92)<br>23.0 (IQR=1.2) |
| AL/CR | 3.0 (SD=0.1)<br>3.0 (IQR= 0.1) | 3.0 (SD=0.1)<br>3.0 (IQR= 0.1) | 3.0 (SD=0.1)<br>3.0 (IQR= 0.1) | 3.0 (SD=0.1)<br>3.0 (IQR= 0.1) |
| Near work (hours/day) | 5.3 (SD=3.1)<br>4.6 (IQR=3.3) | 4.8 (SD=2.6)<br>4.4 (IQR=3.2) | 2.5 (SD=1.6)<br>2.2 (IQR=2.1) | 2.9 (SD=1.8)<br>2.7 (IQR=2.4) |
| Outdoor time (hours/day) | 2.6 (SD=2.2)<br>1.9 (IQR=2.1) | 2.4 (SD=1.7)<br>1.8 (IQR=2.0) | 1.6 (SD=1.0)<br>1.4 (IQR=1.3) | 1.6 (SD=1.2)<br>1.3 (IQR=1.6) |

*SD= standard deviation, IQR= interquartile range, AL/CR = axial length/corneal-curvature ratio, AL = axial length*

**Table 2.**
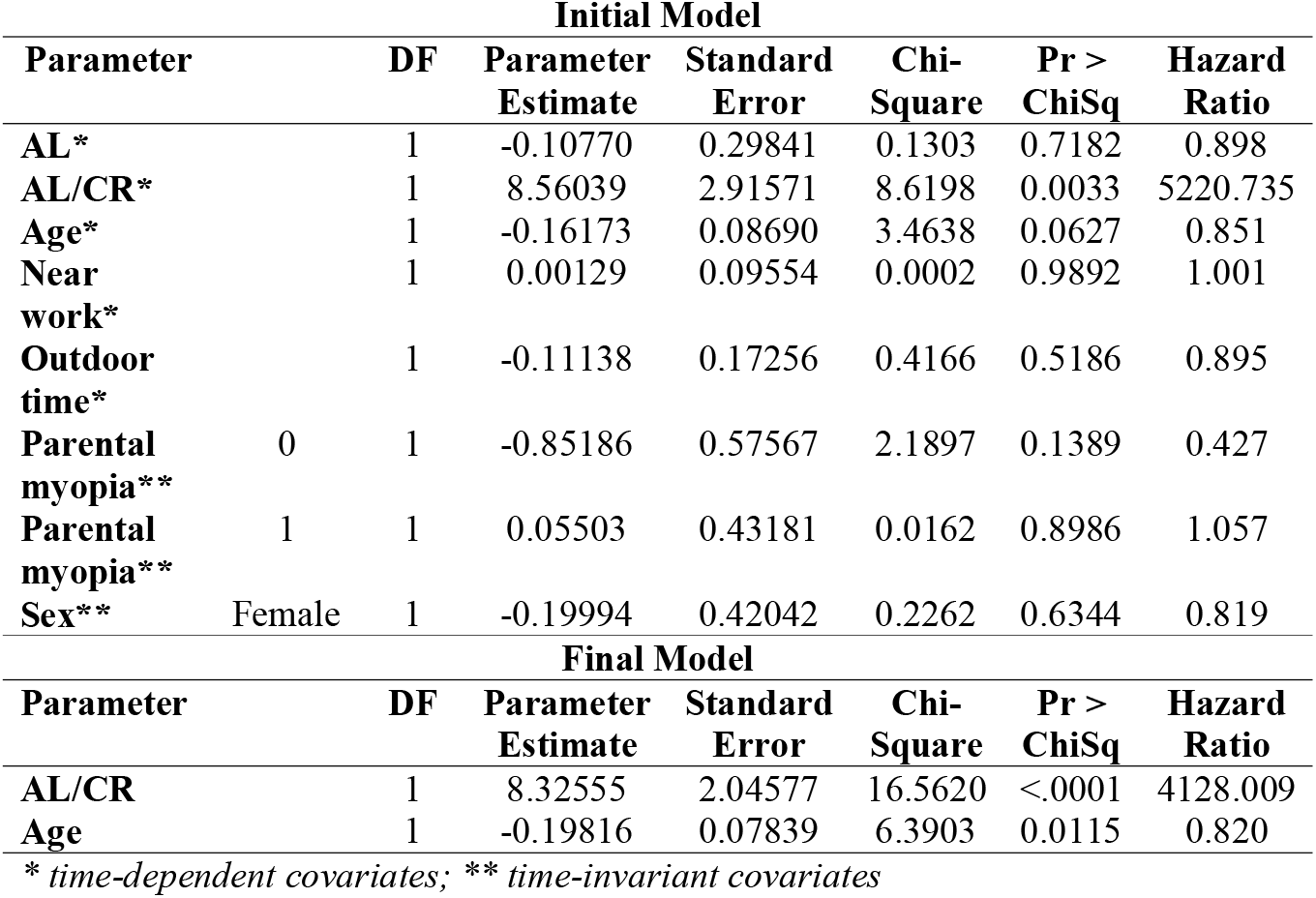
Summary of the Cox proportional hazards regression, at the top the initial model and the bottom the final model. The event modulated was “myopic shift” and multiple events per participant were possible. The null hypothesis was that all regression coefficients in the model are equal to zero. When the p-value (Pr>ChiSq) was less than 0.05 the parameters in the model were considered different from zero, that is, independent predictors of the myopic shift.

The effect of time, ametropia at baseline and parental myopia on SER was tested using linear mixed models in SAS software PROC MIXED (SAS Institute Inc., Cary, NC, USA). Normalized SER was obtained by subtracting the baseline SER from all other measurements, that corresponds to a baseline SER of 0 for all participants, that way it was possible to model SER progression. For this analysis the normalized SER was defined as “dependent variable”. Participants were defined as “random factors”. Explanatory factors or “fixed factors” were: “ametropia” (Myopia, Emmetropia, H**y**peropia), “parental myopia” (0,1,2 myopic parents). Other factors and their interaction with time were tested. P-values were adjusted for multiple comparisons using the Tukey-Kramer procedure. Means in the text and shown in graphs are the estimated means (mean response for each factor, adjusted for any other variables in the model) and their standard errors for the specified factors. Statistical significance was set at p<0.05.

## Results

### Sample characteristics

The cohort was formed of 128 participants, at baseline 70 (54.7%) were females and 58 males (45.3%), during the study seven participants dropped out meaning that 121 participants finished the study, 67 (55.4%) females and 54 (44.6%) males – the retention rate was 95%. The sample was predominantly Caucasian (86.0%), Table 1 summarizes longitudinal values of clinical and sociodemographic variables.

### Incidence and prevalence

At baseline, the prevalence of myopia was 10.0% (CI95=4.4-14.9), hyperopia was 48.0% (CI95= 38.8-56.7), and emmetropia was 42.0% (CI95=33.5-51.2). The *cumulative incidence* of myopia during the two years of follow-up was 5.5% (CI95= 2.2-10.9), and the *incidence rate* of myopia was 3.2 cases per 100 person-years (CI95= 0.8-5.6). The *cumulative incidence* of myopic shift during the two years of follow-up was 21.0% (CI95= 14.4-29.2), and the *incidence rate* of myopic shift was 12.4 cases per 100 person-years (CI95= 7.7-17.1).

### Cox proportional hazards model of myopic shift during follow-up

Table 2 (Initial Model) summarizes the initial Cox regression model with a full list of predictors. AL/CR was the only independent predictor of the risk of myopic shift, age was almost statistically significant. After performing the method of backward elimination, the final model, given in Table 2 (Final Model), included AL/CR and age as predictors of myopic shift.

According to the final model in Table 2, “AL/CR” and Age had a statistically significant effect on the hazard of the event. For each additional unit in age, the hazard of myopic shift decreased by 100*(0.82-1) =18.0%. Hazard increased by 4128 for every 1 unit in AL/CR. In other words, for each additional 0.01 unit in AL/CR ratio, the hazard of having an event of myopic shift increased by: (e^8.3255/100^) -1= 8.7%. Figure 1 shows a survival function for three age category groups.

**Figure 1.**
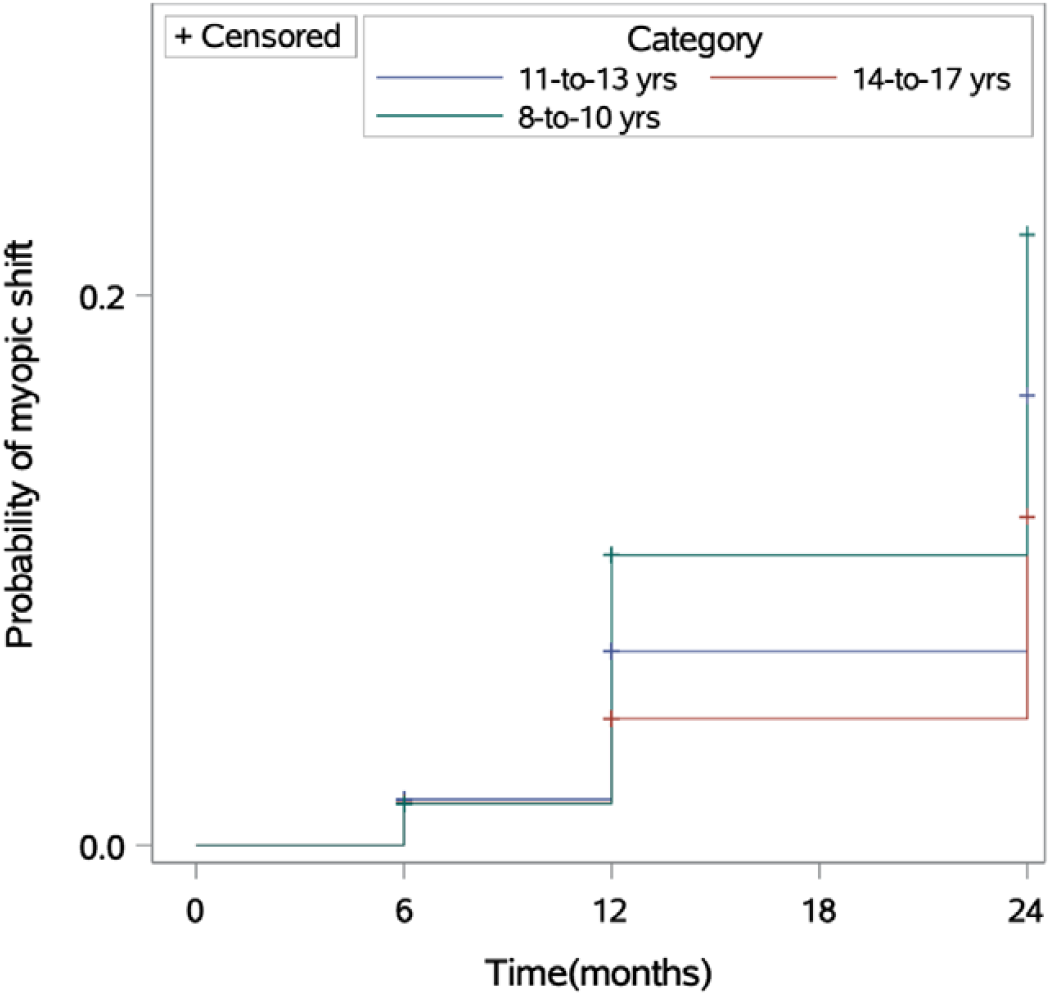
Kaplan-Meier graph assuming AL/CR ratio of a mean value of 2.98. for different age categories. On the y-axis is the probability of having a myopic shift and on the x-axis is the time in months since the start of the study. This graph shows different age categories only to illustrate the effect of age; although, in the Cox regression age was used as a continuous time-dependent covariate.

### Longitudinal changes in SER and associated factors

There was a statistically significant effect of time on SER (F (3, 250) =16.3, p<0.001). Table 3 summarizes pairwise comparisons with adjusted p-values of SER between different time points.

**Table 3.** Summary of pairwise comparisons of SER between time points: baseline (0), 0.5, 1, and 2 years. The mean difference (M) was computed by subtracting SER at baseline in the column from SER in the row. SE= standard error (SE), p-values were adjusted with Bonferroni correction for multiple comparisons.

| Time | 0 | 0.5 | 1 |
| --- | --- | --- | --- |
| 0.5 | M= 0.004 D (SE=0.04),<br>p=0.063 | - | - |
| 1 | M= -0.16 D (SE= 0.05),<br>p=0.005 | M= -0.15 D (SE= 0.05),<br>p=0.006 | - |
| 2 | M= -0.27 D (SE= 0.04),<br>p<0.001 | M= -0.26 D (SE= 0.04),<br>p<0.001 | M= -0.11 D (SE= 0.04),<br>p=0.063 |

There was a significant interaction ametropia×stime (F (2, 122) = 6.36, p=0.002), these results are summarized in Table 4 and Figure 2-A. There was a significant interaction time×parental myopia (F (2, 122) = 6.16, p=0.003), these results are summarized in Table 4 and Figure 2-B.

**Table 4.** Summary of pairwise comparisons of SER changes between categories of ametropia on the left-hand side and parental myopia on the right-hand side at time points: baseline (0), 0.5, 1, and 2 years. The mean difference (M) was computed by subtracting SER for the time in the column “Time” from SER for the time in the first row. SE= standard error, p-values were adjusted for multiple comparisons.

| Ametropia | Time | 0 | 0.5 | 1 | Myopic parents | Time | 0 | 0.5 | 1 |
| --- | --- | --- | --- | --- | --- | --- | --- | --- | --- |
| Myopia | 0.5 | M= -0.10 D<br>(SE= 0.10),<br>p=0.286 | - | - | none | 0.5 | M= -0.03 D<br>(SE= 0.07),<br>p=1.00 | - | - |
|  | 1 | **M= -0.30 D**<br>(SE= 0.12),<br>**p=0.011** | M= -0.20 D<br>(SE= 0.12),<br>p=0.085 | - |  | 1 | M= 0.02 D<br>(SE= 0.10),<br>p=1.00 | M= 0.05 D<br>(SE= 0.01),<br>p=1.00 | - |
|  | 2 | **M= -0.56 D**<br>(SE= 0.11),<br>**p<0.001** | **M= -0.46 D**<br>(SE= 0.10),<br>**p<0.001** | M= -0.26 D<br>(SE= 0.11),<br>p=0.365 |  | 2 | M= -0.11 D<br>(SE= 0.08),<br>p=0.967 | M= -0.10 D<br>(SE= 0.07),<br>p=0.993 | M= -0.13 D<br>(SE= 0.07),<br>p=0.863 |
| Hyperopia | 0.5 | M= 0.004 D<br>(SE= 0.05),<br>p=0.947 | - | - | 1 | 0.5 | M= 0.11 D<br>(SE= 0.06),<br>p=0.866 | - | - |
|  | 1 | M= -0.10 D<br>(SE= 0.06),<br>p=0.931 | M= -0.10 D<br>(SE= 0.06),<br>p=0.946 | - |  | 1 | M= -0.14 D<br>(SE= 0.07),<br>p=0.746 | **M= -0.24 D**<br>(SE= 0.07),<br>**p=0.029** | - |
|  | 2 | M= -0.16 D<br>(SE= 0.06),<br>p=0.240 | M= -0.16 D<br>(SE= 0.05),<br>p=0.153 | M= -0.07 D<br>(SE=0.06),<br>p=0.989 |  | 2 | M= -0.23 D<br>(SE= 0.07),<br>p=0.075 | **M= -0.33 D**<br>(SE= 0.07),<br>**p<0.001** | M= -0.09 D<br>(SE= 0.07),<br>p=0.960 |
| Emmetropia | 0.5 | **M= 0.09 D**<br>(SE= 0.05),<br>**p=0.046** | - | - | 2 | 0.5 | M= -0.10 D<br>(SE= 0.08),<br>p=0.991 | - | - |
|  | 1 | M= -0.08 D<br>(SE= 0.05),<br>p=0.941 | **M= -0.18 D**<br>(SE= 0.05),<br>**p= 0.001** | - |  | 1 | **M= -0.37 D**<br>(SE= 0.10),<br>**p=0.006** | M= -0.28 D<br>(SE= 0.10),<br>p=0.110 | - |
|  | 2 | M= -0.08 D<br>(SE= 0.05),<br>p=0.956 | **M= -0.17 D**<br>(SE= 0.05),<br>**p=0.020** | M= -0.01 D<br>(SE= 0.05),<br>p=1.00 |  | 2 | **M= -0.47 D**<br>(SE= 0.09),<br>**p<0.001** | **M= -0.38 D**<br>(SE= 0.08),<br>**p<0.001** | M= -0.10 D<br>(SE= 0.08),<br>p=0.986 |

**Figure 2.**
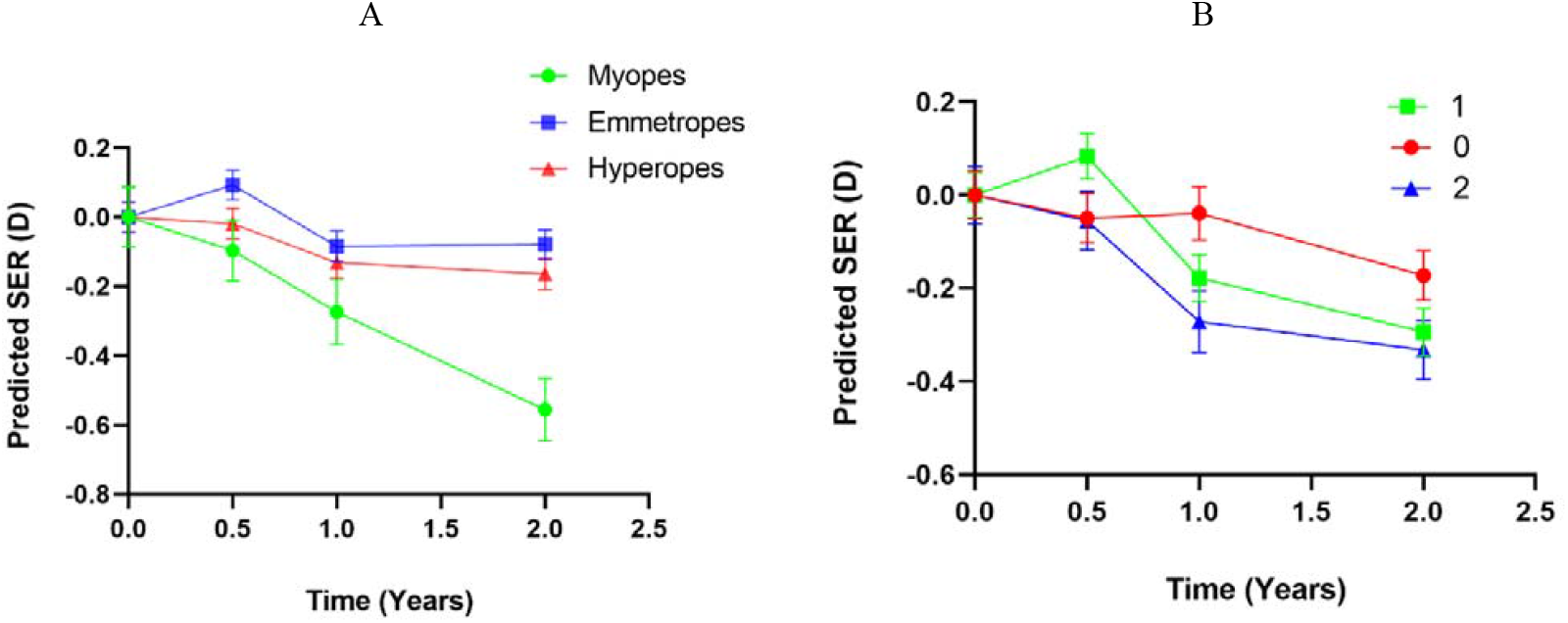
Multiple line graph showing results of the linear mixed model analysis. On the y-axis is the predicted change in SER and on the x-axis is the time in years with measurements performed at: baseline (0 years), 0.5 years, 1 year, 2 years. A) Progression of SER according to ametropia at baseline. B) Progression of SER according to parental myopia (none, one, or two myopic parents). Means in graphs are the estimated means (mean response for each factor, adjusted for any other variables in the model) and error bars represent the standard error of the mean.

## Discussion

This is the first longitudinal study conducted in Scandinavia investigating the incidence of myopia and the incidence of myopic shift in a sample of schoolchildren. The study also investigated factors associated with myopia development and progression. Age and AL/CR showed to be independent predictors of the risk of myopic shift. Ametropia at baseline and parental myopia showed to be factors affecting progression of SER over time. When we compared SER changes between visits in Table 3, there was no statistically significant SER change between visit 1 and visit 2, where visit 2 was during the peak of the pandemic. This points to a reduced effect of the pandemic restrictions in our sample. Although, it must be noted that restrictions in Sweden were few with no complete lockdowns compared, for example, with countries like China that have been included in a recent systematic review and meta-analysis. The review points to a detrimental effect of the pandemic with faster than normal development of myopia^13^.

The two-year cumulative incidence of myopia and the incidence rate of myopia in the current study was consistent with the low baseline prevalence of myopia^3^. The findings of this study are in line with studies from other Scandinavian countries^1,2,14-16^. A study from Denmark compared the myopia prevalence in 29 Danish studies between the years 1882 and 2018 and found no evidence of an increase in myopia numbers over the years^1^. In contrast with our findings, in East Asian countries the cumulative incidence of myopia ranges between 33.6% and 54%^17^, and the annual incidence rate of myopia varies from 30 cases per 100-person years^18^ to 31.7 cases per 100-person years^19^. The incidence of myopia tends to be higher in children with East Asian ethnicity than in European Caucasians. It is known that, for example, the prevalence of myopia is higher amongst British South Asian children than amongst British Caucasian children despite their exposure to the same environment^20^. Overall, the incidence of myopia in the current study is lower than in East Asia and is in line with the expectations for Scandinavian populations.

The current study is the first to report the incidence of the myopic shift in Swedish or in Scandinavian children. Given the originality of our findings we can only compare them with other parts of the globe. Studies from East Asia have investigated the one-year cumulative incidence of myopic shift which varied between 51% and 74.6%^19,21,22^. The incidence for myopia and myopic shift in our study were remarkably low compared with parts of the global where myopia is also more prevalent. It remains to be understood which factors explain these differences, but ethnicity, environmental factors and their interaction are expected to be relevant^23^.

The Cox-regression model revealed that age and AL/CR were independent predictors of myopic shift. The results indicated that age is expected to reduce the probability of myopic shift, that is, younger children were at a greater risk of having a myopic shift. These findings are in line with other studies showing that younger age is associated with a faster progression of myopia when compared to older ages^24^. Studies show a decline in progression speed with increasing age in young myopes of both European and Asian ethnicity^25-27^. Our results for age are in line with the current notion that it is important to delay the onset of myopia to prevent high myopia later in life^28^. An increase in the AL/CR ratio increased the probability of myopic shift. The AL/CR ratio has a typical value of 3.0 in emmetropic eyes and more than 3.0 in myopic eyes^29,30^. Our results are in line with other studies showing that increasing AL/CR ratio is associated with a more myopic refractive error^29,30^. These finding are interesting and deserve further investigation because recent studies have shown that AL/CR ratio explains the total variance in refractive error better than AL alone and is an influential variable for myopia detection in children^31,32^. Our results for age give further evidence that is important to delay the onset of myopia^28^. For AL/CR, our findings are consistent with the often-successful results of orthokeratology for myopia management^33^, that is, increasing (flattening) the CR leads to reduction of myopia^33^.

The longitudinal analysis of changes in SER showed the expected trend in children, that is, a change towards negative values. The progression was influenced by ametropia and parental myopia. In the current study the mean change in SER was -0.30 D, that is approximately -0.15 D per year. Reports from East Asia show a typical progression - 0.60 D per year^34,35^ and that is 4-times our progression. These results explain the low incidence of myopia in our study when compared to East Asia. Our findings showed that participants with myopia at the baseline had a significant faster SER change during the study. This results are in line with a recent study reporting myopia progression in European children with myopia^15^. Another independent factor associated with progression of SER was parental myopia. In line with previous studies, children with two myopic parents had a faster change towards myopia than the other two categories^16,36^.

In our cohort the incidence and progression of myopia was low and this raises the possibility that other factors such as high educational pressure can be associated with the high prevalence and progression of myopia in East Asian countries^37^. Several recent studies have concluded that exposure to education in early primary school years with its study pressures and limitations on time outdoors is the major determinant of myopic shift^37,38^. Giving China as an example, there is a strict policy for enrolment in primary schools, where children who have turned six years of age are qualified to start school. Chinese primary school children attend school for seven hours a day^39^. In Sweden, children start primary school at age seven and attend school on average 4.5 hours a day. Also, in the early years of education, schools in Sweden tend to promote outdoor educational activities and long breaks between teaching sessions^40^. Therefore, we speculate that if other follow the Swedish educational curriculum maybe the incidence of myopia will reduce^41^.

The current study has two major strengths, the first strength was its longitudinal design and the second was the low dropout rate. A possible limitation was the poor recall of the information reported in the questionnaire. Even though, some studies have shown that key answers tend to be reasonably accurate^42,43^. Another possible limitation was the sampling method. Since the participants had to travel to the facilities of the faculty where the study was based, it is unlikely that the sample was a random selection of the population. This might have attracted overly concerned children and parents only, but given the low myopia prevalence, this limitation seems to have had limited impact on the findings. Further, the sample size limits the statistical power to find associations between environmental factors and myopia. However, our findings are in line with the most recent suggestions that environmental conditions such as educational stress at a very young age may impact myopia onset and progression^37^. Further studies are warranted for investigating the effect of educational pressure on refractive error development amongst Swedish children.

## Conclusion

In this longitudinal study with a cohort formed mostly by Caucasian children living in Sweden. The incidence of myopia and myopic shift was low when compared with other parts of the globe. Younger children were at a greater risk of having a myopic shift. Myopic children at the baseline and children with two myopic parents showed a significant faster-paced refractive error change over time. Our results show that parental myopia remains a critical confounder to consider when planning clinical trials for myopia control interventions. Further studies are necessary involving children from different ethnicities living in Sweden under the same environmental conditions to investigate myopia prevalence and myopia progression patterns.

## Data Availability

All data produced in the present study are available upon reasonable request to the authors

## Acknowledgments

This study was supported by Specsavers Sweden AB, the faculty of Health and Life Sciences, Linnaeus University, and Brien Holden Vision Institute.

